# Extending the Ecological Model of Distress to Social Functioning among Refugees and Asylum-seekers

**DOI:** 10.64898/2025.12.09.25341945

**Authors:** Gulsah Kurt, Philippa Specker, Belinda Liddell, David Keegan, Randy Nandyatama, Atika Yuanita, Rizka Argadianti Rachmah, Joel Hoffman, Angela Nickerson

## Abstract

**Aim:** Social functioning is a crucial aspect of psychosocial adaptation following forced displacement. Yet, it has received far less attention than understanding and addressing mental health problems among refugees and asylum-seekers. This study aimed to extend the ecological model of distress - one of the most widely used frameworks in refugee mental health - to social functioning, and to identify direct and indirect pathways from established conflict- and displacement-related factors to social functioning alongside mental health problems.

**Method:** An online study with 1,235 refugees in Indonesia was conducted over a two-year period. Conflict-related traumatic experiences before arrival in Indonesia, post-displacement stressors in the past 12 months before baseline, were measured at the onset of the study, while social functioning and mental health outcomes (symptoms of posttraumatic stress disorder, depression, and anger) were assessed one year later.

**Results:** Longitudinal Structural Equation Modelling analysis revealed that pre-migration trauma predicted more post-displacement stress, higher mental health problems, but increased social functioning one year later, while post-displacement stressors predicted poorer mental health and reduced social functioning. The indirect pathway from traumatic experiences via post-displacement stressors was positive for mental health and negative for social functioning.

**Conclusions:** This study conceptually and empirically extended the ecological model of distress to social functioning among refugees by highlighting the dual influences of conflict-related traumatic experiences. The findings provide a springboard for advancing research and practice in the mental health and psychosocial field.

## Introduction

The number of forcibly displaced people continues to rise, with 45 million refugees and asylum-seekers worldwide. These people experience prolonged human rights violations, exposure to multiple traumatic events, and persistent daily stressors, as documented across diverse contexts and over decades (Hou et al., 2020; Sisenop et al., 2025; Steel et al., 2009). Such experiences increase the risk of developing mental health problems (Blackmore et al., 2020). Forced displacement also erodes the social fabric of societies and disrupts social networks, relationships, and resources (Ager & Strang, 2008; Silove, 2013; Strang & Quinn, 2021). Despite the importance of these social sequela, research and practice among those forcibly displaced have largely focused on mental health outcomes, with limited attention to social functioning as a central component of wellbeing and psychosocial adaptation.

Several theoretical models have been developed to elucidate the link between stressors related to conflict and forced displacement, and the mental health of forcibly displaced populations. Among these, the ecological model of refugee distress developed by Miller and Rasmussen has gained increasing attention since its initial introduction in 2010 (Miller & Rasmussen, 2010) and subsequent iteration in 2017 (Miller & Rasmussen, 2017). This model posits that both conflict-related traumatic events and socio-environmental stressors following post-conflict and forced-displacement adversely impact mental health. As conflict and war often cause or exacerbate socio-environmental stressors encountered in daily life, these experiences exert their influence on mental health both directly and indirectly through these stressors. Given its simplicity and explanatory power, the ecological model has been the focus of considerable scientific inquiry over the past decade (Miller & Rasmussen, 2024) and has received substantial empirical support (e.g., Dangmann et al., 2021; EL-Awad et al., 2025; Goodkind et al., 2021; Hou et al., 2020). Studies consistently demonstrated that both conflict-related traumatic events and stressors in post-displacement settings, whether in resettlement contexts or during transit, significantly predict increased mental health problems, with post-displacement stressors showing overall stronger association than traumatic experiences (Hou et al., 2020). Although the later version of the model has been expanded to family functioning, such as parenting and parental mental health, it remained largely focused on understanding the predictors and consequences of poor mental health among conflict-affected populations. While this model has substantially advanced our understanding of conflict- and displacement-driven mental health problems, it represents only one aspect of psychosocial adaptation following such experiences.

Social functioning - the ability to form and maintain meaningful relationships and engage in social activities (Bosc, 2000; Hirschfeld et al., 2000) is an important yet often underexplored dimension of adaptation and wellbeing in conflict-affected populations (Ager & Strang, 2008; Lahiri et al., 2017). Social functioning is closely related to mental health, as mental health difficulties are often accompanied by impairments in social functioning in both general (Scoglio et al., 2022) and conflict-affected populations (Kurt et al., 2023; Nickerson et al., 2025a). Yet, social functioning represents a distinct construct that extends beyond mental health symptoms, and difficulties in social functioning often persist even after mental health symptoms have subsided (Kupferberg & Hasler, 2023; Scoglio et al., 2022). Improved social functioning, is often considered among the key recovery goals following adversities and mental health problems (Guerrero et al., 2024; Vera San Juan et al., 2021). For conflict-affected people, especially for refugees and asylum-seekers, social functioning is key to successful integration and psychosocial adaptation (Strang & Quinn, 2021). However, existing evidence on social functioning is scant, with most research relying on cross-sectional designs. A recent systematic review of 38 studies found that the majority of evidence supports the adverse impact of conflict-related traumatic events on social functioning, but some studies show that these experiences might also lead to increased social functioning outcomes, including greater participation in community and cultural groups and more frequent interaction with others (Perkins et al., 2025). Further, other studies indicated that stressors encountered in forced displacement contexts likely impair social functioning (Nguyen et al., 2024; J. L. Steel et al., 2017; Wachter et al., 2016). Despite evidence supporting the role of both conflict- and displacement-related factors for social functioning, no study to date has examined these factors together through the proposed direct and indirect pathways in Miller and Rasmussen’s model (Miller & Rasmussen, 2017), in relation to social functioning within the same model as mental health.

The present study aimed to longitudinally test the ecological model of distress in relation to social functioning among refugees and asylum seekers in Indonesia, to identify similarities and differences between the determinants of mental health and social functioning. Consistent with the ecological model, we hypothesized that there would be direct pathways from conflict-related traumatic events and post-displacement stressors to mental health outcomes as well as an indirect pathway from traumatic events to mental health via post-displacement stressors. Given emerging but still limited evidence on the potential dual role of conflict-related traumatic experiences for social functioning, we explored whether there would be a direct pathway from conflict-related traumatic events to social functioning without specifying the direction. Drawing from the established literature, we hypothesized that traumatic events would predict greater post-displacement stressors, which would in turn predict poorer social functioning. The extended ecological model for social functioning is depicted in Figure 1.

**Figure 1.**
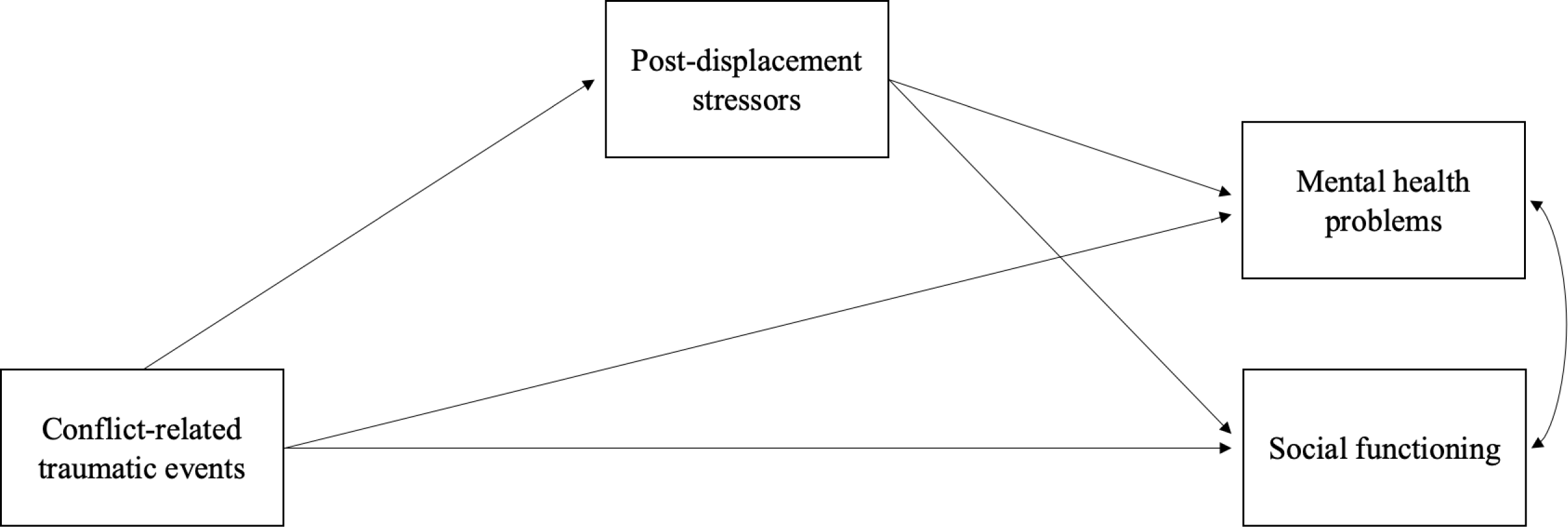
The extended ecological model of distress for social functioning.

## Methods

### Participants and Study Design

The present study used data from first and third waves of a large-scale longitudinal study conducted with refugees and asylum-seekers in Indonesia between 2020 and 2022. The first wave (Time 1) was collected between February and October 2020, and the third wave (Time 3) was collected 12 months after the completion of Time 1. Participants were recruited through refugee services, community-based organizations in Jakarta and Bogor, Indonesia, and social media. Eligibility criteria were: (1) being a refugee or asylum seeker in Indonesia, (2) having arrived in Indonesia after January 2013, (3) being at least 18 years old, and (4) being literate in one of the following languages representing the majority of refugees at the time of the study: Arabic, Farsi, Dari, Somali, or English. Participants completed a self-administered online survey (approximately 1 hour) at each wave. The trained research assistants provided technical support when needed. Participants were compensated with an online grocery voucher of USD 7 (IDR 100,000) for their participation. Ethical approval was obtained from the UNSW Human Research Ethics Committee (HC190494) and Atma Jaya University, Jakarta (0792/III/LPPM-PM.10.05/07/2019).

### Measures

All measures were translated and blind back-translated following a rigorous translation process by accredited translators. Refugee community members with varying levels of education pilot-tested the translated survey battery to assess comprehension and clarity.

### Conflict-related traumatic events

Items from the Harvard Trauma Questionnaire (16 items; Mollica et al., 1992) with additional three items (natural disaster, physical assault, and serious accident, fire or explosion) were used to assess potentially traumatic experiences related to war and conflict that participants had either directly experienced or witnessed in the past, using categorical response options (0 = No, 1 = Yes). The total number of endorsed traumatic events (experienced and/or witnessed) was used to represent diversity of conflict-related traumatic experiences in this study.

### Post-displacement stressors

An adapted version of the Post-migration Living Difficulties Checklist (Z. Steel et al., 1999) was used to measure the stressors that participants experienced in their daily lives in Indonesia in the past 12 months. 42 items were presented to the participants to rate on a 5-point Likert Scale (1= *was not problem/did not happen,* 5= *a very serious problem)*. The mean score of the overall items was used in the study. The Cronbach’s alpha was 0.96 in the present study.

### Mental health outcomes

*Depression symptoms* were measured using the Patient Health Questionnaire-8 items (PHQ-8) (Kroenke et al., 2009). Participants indicated how much they had been bothered by each item on a 4-point Likert scale (1 = *not at all,* 4 = n*early every day*). The mean score across items was used in this study (α = 0.93). An adapted version of the Posttraumatic Diagnostic Scale for DSM-IV (Foa et al., 1997) was used to measure the symptoms of posttraumatic stress disorder. Four items reflecting the revised PTSD symptoms in DSW-5 was added to the original 16 items. Participants were asked to report how often each PTSD symptom bothered them over the past month on a 4-point Likert Scale (0= *not at all/only once*, 3= *5+times a week/almost always*). The mean score across 20 items was used in this study (α= 0.96). Anger symptoms were measured with the 5-item Dimensions of Anger Reactions Scale (Forbes et al., 2014) rated on a 5-point Likert Scale (1= *none or almost none of the time*, 5= *all or almost all of the time)*. The mean score of the items was used in this study (α= 0.92).

### Social functioning

Social functioning was assessed using three items from the Short Social Capital Assessment Tool (De Silva et al., 2007). These items covered different behavioral aspects of social functioning: *Active group membership* was indexed by the number of groups in which participants were active members over the past 12 months, out of seven group types (community groups, women’s groups, men’s groups, religious groups, sports groups, student groups, volunteer/charity groups). *Support from groups* was indexed by the number of groups from which participants received emotional or economic assistance over the past 12 months, out of 12 groups (community groups, women’s groups, men’s groups, religious groups, sports groups, student groups, volunteer/charity groups, charitable organizations, refugees from own community and from other communities and members of Indonesian community) *Support from individuals* was indexed by the number of types of individuals from whom participants received emotional or economic assistance over the past 12 months, out of 14 categories (family, neighbors, friends, community leaders, politicians, religious leaders, government officials, charitable organizations, volunteers, refugees from one’s own community, refugees from other communities, members of the Indonesian community, non-refugees from other communities, and advocates).

Each item represents a count variable indicating the number of categories endorsed by the participants. A total social functioning score was calculated by summing these items (Nickerson et al., 2022).

### Demographic variables

Key demographic information, such as age, gender, language, time spent in Indonesia, separating from family (having no family member vs. all or some family member in Indonesia) was collected.

## Data Analysis

We used longitudinal structural equation modeling in Mplus Version 8.2 (Muthen, L.K & Muthen, B.O, 2023) to test the associations between conflict-related traumatic experiences, post-displacement stressors and mental health and social functioning outcomes. Following a two-stepped approach, we first conducted Confirmatory Factor Analysis (CFA) using the mean scores of the symptoms of PTSD, depression, and anger at T3 to form a latent variable of mental health problems. Then, we tested our hypothesized mediation model using conflict-related traumatic events before arrival in Indonesia measured at T1, post-displacement stressors in the past 12 months measured at T1, latent variable of mental health problems at T3, and observed variable of social functioning at T3. Social functioning was treated as an observed variable because it comprises behaviorally anchored indicators that reflect directly observable actions rather than an underlying latent construct. We estimated direct paths from conflict-related traumatic events and post-displacement stressors to mental health and social functioning as well as indirect paths from traumatic experiences via post-displacement stressors to these outcomes. The significance of the indirect predictor role of conflict-related traumatic events on mental health and social functioning outcomes via post-displacement stressors was tested using the bootstrapping technique (1000 resampling) (MacKinnon et al., 2007). The model controlled for age, gender, separation from family, time in Indonesia, and language groups (as a proxy for country of origin). Model fit was evaluated using the following indices: Comparative Fit Index (CFI) >.90, Root Mean Square Error of Approximation (RMSEA) <.08 with 90% CI and Standardized Root Mean Square Residual (SRMR) <.08 (Brown, 2015; Kline, 2023; MacCallum et al., 1996). Full information maximum likelihood estimation was used to account for missing data on endogenous variables. The missing rate was less than 5% on the exogenous variables; therefore, no imputation was conducted.

## Results

### Participants

A total of 1235 participants completed Time 1 with an average age of 30.53 (SD= 9.067). The majority of the sample was male (71.4%) and were from Farsi/Dari speaking backgrounds (39.4%) followed by Arabic (30.3%), English (17.5%), and Somali (12.9%). The average length of stay in Indonesia was 5.11 years (SD=1.618). Half of the participants (53.5%) reported having no immediate family member in Indonesia. The average number of conflict-related traumatic experiences as 7.73 (SD=5.03) with lack of food or water (54.1%), ill health without access to healthcare (51.8%), and lack of shelter (42.5%) reported as the most common experiences. The mean of post-displacement stressors was 2.81 (SD= 0.86), with the most common issues being difficulties related to work, fear of being sent back to the home country, and worries about visa status.

### Measurement Model

CFA yielded an overall good model fit for mental health as a latent variable (CFI= 1.00, RMSEA= 0.01, SRMR= 0.02) indexed by mean symptoms of PTSD, depression, and anger. The model was just identified with factor loadings of 0.89 for depression, 0.90 for PTSD, and 0.80 for anger, indicating acceptable loadings.

### Structural Equation Model Testing

#### Direct paths from conflict-related traumatic events and post-displacement stressors to mental health and social functioning

The model fit the data well, CFI=0.98, TLI=0.95, RMSEA=0.04 (90% CI= 0.023-0.047), SRMR=0.02. Figure 2 depicts significant standardized direct effects. Conflict-related traumatic events before arrival in Indonesia predicted greater post-displacement stressors at T1 (β= 0.45, SE= 0.03, *p <*.001). Conflict-related traumatic events and post-displacement stressors at T1 predicted greater mental health symptoms at T3 (β=0.13, SE=0.05, *p* = 0.004, β=0.46, SE=0.05, *p* <.001). Conflict-related traumatic events predicted better social functioning at T3 (β= 0.10, SE= 0.04, *p* = 0.011) while post-displacement stressors at T1 predicted worse social functioning at T3 (β= -0.09, SE= 0.04, *p*= 0.041).

**Figure 2.**
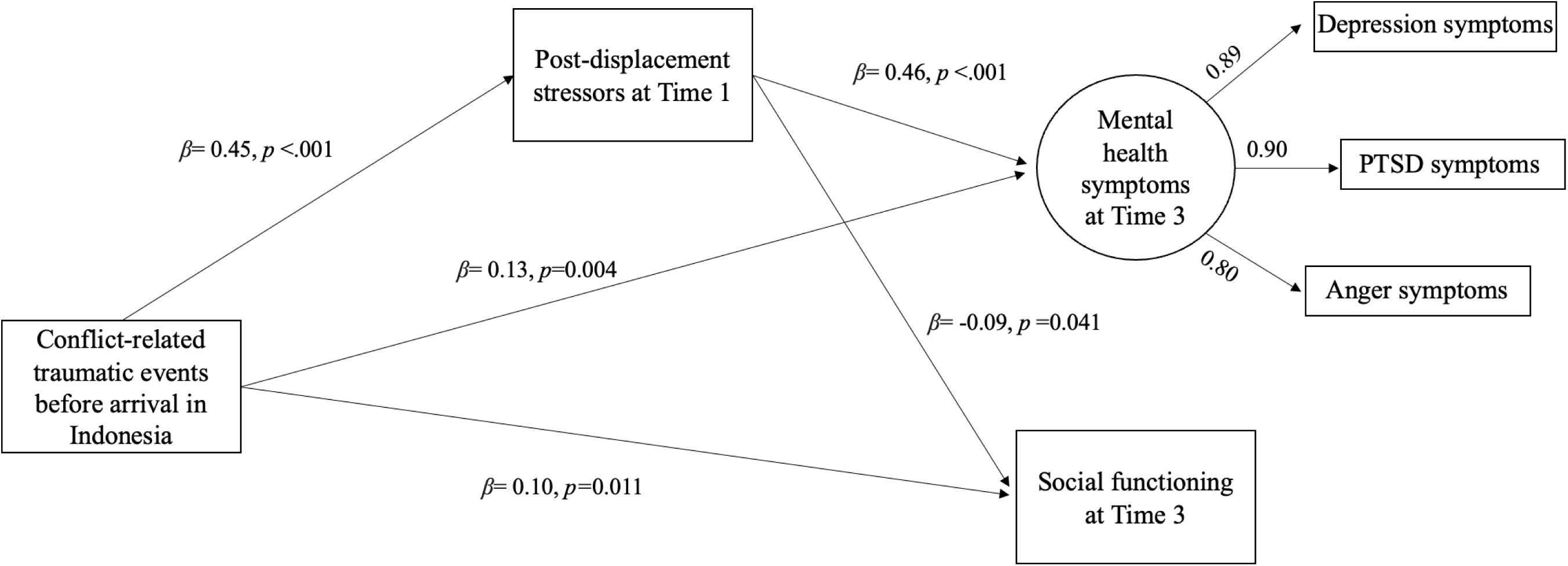
The extended ecological model for refugee distress to social functioning.

#### Indirect path from conflict-related traumatic events to the outcome variables via post-displacement stressors

The indirect associations of conflict-related traumatic events on mental health problems and social functioning via post-displacement stressors were significant (β= 0.21, 95%CI =0.162 to 0.257, β= -0.04, 95%CI= -0.082 to -0.003). That is, conflict-related traumatic events predicted greater post-displacement stressors at T1, which, in turn, predicted greater mental health symptoms and worse social functioning at T3. Table 1 includes the full list of standardized coefficients for direct and indirect paths.

**Table 1.**
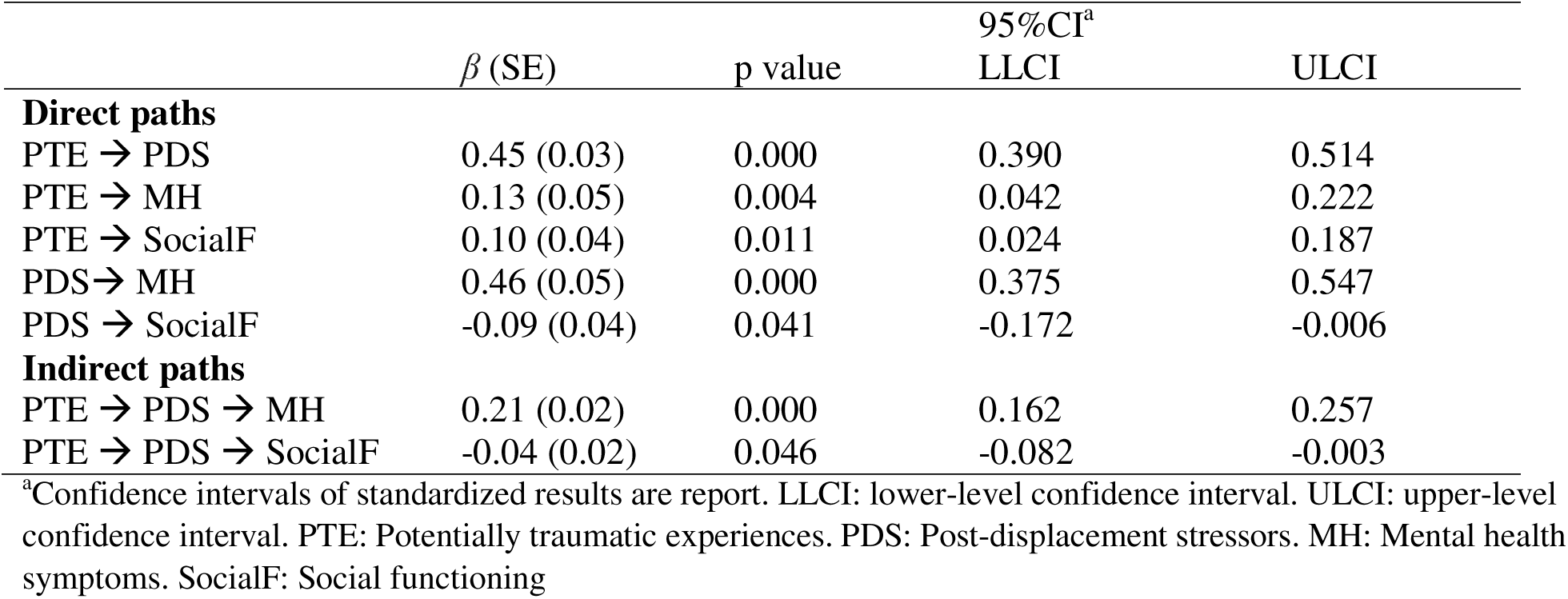
Standardized direct and indirect paths.

## Discussion

In the present study, we longitudinally tested an extension of the ecological model of distress to social functioning in a sample of culturally and linguistically diverse refugees and asylum seekers in Indonesia. Our results accord with the established pathways from conflict-related traumatic experiences and post-displacement stressors to mental health, while highlighting both shared and unique associations for social functioning. The findings provide the first empirical evidence that social functioning is distinctly predicted by conflict- and displacement-related stressors.

Consistent with prior research (Z. Steel et al., 2009) and our hypotheses, we found that conflict-related traumatic experiences predicted greater mental health problems via both direct and indirect pathways. These experiences were associated with greater post-displacement stressors in the past year, which, in turn, predicted higher levels of mental health problems one year later. The observed longitudinal pathways for mental health align with the accumulated evidence for the ecological model of distress and contribute to the relatively sparse longitudinal research in this area (Miller & Rasmussen, 2024). Further, the results revealed a dual pathway from conflict-related traumatic events to social functioning. That is, experiencing conflict and war-related traumatic events before arrival in Indonesia (such as imprisonment, loss of loved ones, and deprivation of basic needs) was directly associated with greater social functioning, operationalized as greater engagement with and support from groups and individuals. In contrast, exposure to these traumatic events was also indirectly associated with worse social functioning over time via greater post-displacement stressors. These findings are particularly novel as they suggest that conflict-related traumatic experiences can have dual influences on social functioning, largely shaped by the post-displacement context.

The dual pathway from conflict-related traumatic events to social functioning might reflect distinct underlying processes. Regarding the pathway by which trauma exposure is associated with greater social functioning, experiencing traumatic events such as losing loved ones may motivate individuals to interact more with others and seek resources and support to restore connection disrupted by trauma (Matos et al., 2021; Posselt et al., 2019). To do so, they might engage in social activities, such as participating in social and community groups and seeking social support from other individuals and the groups that they are part of (Frounfelker et al., 2020; Nickerson et al., 2019). Emerging evidence supports this notion, showing that experiences of conflict-related events can be associated with increased participation in social groups, greater support from close others, and more engagement with host culture (Perkins et al., 2025). At the same time, the indirect pathway whereby the same conflict-related traumatic events predicted poorer social functioning through higher post-displacement stressors indicates the possibility of more than one behavioral route, shaped by the degree or type of contextual stress. One potential direct pathway may occur via increased motivation to approach others, while a second pathway may occur through the accumulation of post-displacement stressors that constrain behavioral engagement in social activities. This is consistent with the physiology of stress regulation, which describes the stress response as producing both mobilizing (approach) and hindering (avoidance) effects that operate non-linearly depending on the severity and duration of stress exposure to maintain equilibrium between stress exposure and one’s coping capacity (Gray, 1984; Kasch et al., 2002; McEwen, 2007).Thus, in our case, conflict-related trauma might activate behavioral approach systems to restore needs for connection and belonging, while greater chronic exposure to post-displacement stressors might engage inhibitory processes that reduce social engagement. These parallel processes can explain the dual pattern of associations with trauma observed in our study.

It is important to interpret the current findings within the context in which this study was conducted. In Indonesia, asylum-seeking claims are only processed by UNHCR as the country is not a signatory to the Refugee Convention and its protocol. This often means that individuals seeking asylum must wait for years without access to formal employment and limited access to health care and education (Amin, 2022; Curby, N, 2020). Thus, systemic injustices, insecurity, and financial hardships pose significant structural barriers that preclude these individuals’ meaningful participation in social life (Ager & Strang, 2008; Niemi et al., 2019). In the absence of supportive legal and social structures, refugees and asylum seekers are presented with limited opportunities to engage in social activities (Niemi et al., 2019). This situation portrays the structural barriers and systemic exclusion of refugees from participation common across transit, low-resource settings (Hynie, 2018). Addressing post-displacement stressors is therefore not only critical for mental health but also for fostering social functioning.

The study findings should be interpreted within the context of some limitations. Firstly, we focused on the behavioral dimension of social functioning, indexed by group memberships and the support received from social networks. Although these behavioral indicators have been consistently linked to health and functioning benefits, especially after significant life transitions (Cruwys et al., 2013; Haslam et al., 2021; Iyer et al., 2009; Stephen et al., 2021), they do not necessarily convey any valence (positive or negative) regarding the quality or satisfaction of one’s social functioning. These dimensions are equally important to behavioral indicators to capture a comprehensive understanding of social functioning as a phenomenon. Thus, future studies should measure these and other additional indicators of social functioning. Another limitation is online nature of data collection which enabled us access to usually hard-to-reach communities in our study. However, this may have resulted in more participation from individuals with higher digital literacy. We also used non-random sampling, which might limit the generalizability of the findings to the broader refugee population in Indonesia and beyond. Nonetheless, the large sample size and the similarity of its demographic characteristics to those of the target population at the onset of the study (United Nations High Commissioner for Refugees, 2018) support the potential validity of our findings among refugees in Indonesia.

Despite these limitations, the present study has several implications for research, practice, and policy to improve overall refugee wellbeing and psychosocial adaptation. First, our findings contribute to the burgeoning research highlighting the distinct nature of social functioning relative to mental health among refugees and asylum seekers (Kurt et al., 2023; Nickerson et al., 2025b). For the first time, this study provides empirical evidence supporting the extension of the commonly used ecological model of distress to social functioning and illustrates the distinct pathways linking established risk factors for mental health to social functioning. These findings lay the groundwork for future research to validate similar patterns of association with diverse refugee populations in different transit and resettlement contexts. Given the differential associations of conflict-related traumatic experiences and post-displacement stressors, examining mechanisms underpinning these associations for mental health and social functioning is necessary. Further research is also warranted to investigate the granularity of social functioning by focusing on multiple domains of social life. In this regard, it may be beneficial to draw on approaches previously used to understand cultural meaning and expressions of mental health (e.g., Keys et al., 2012; Kirmayer, 2025; Ventevogel et al., 2013; Ventevogel & Faiz, 2018). Similar to any other construct, social functioning is socially constructed - shaped by cultural and societal values, norms, and expectations. These cultural and societal factors likely determine what it means to be socially functioning well. Understanding these influences and the cultural meaning of social functioning, then, becomes crucial for defining the construct accurately, measuring it appropriately, and designing effective strategies and interventions to promote it (Greene et al., 2023; Miller et al., 2006; Ubels et al., 2022).

Considering the role of conflict-related traumatic experiences and post-displacement stressors in both mental health and social functioning, our findings contribute to the call for multilevel interventions that address trauma-related difficulties while simultaneously improving socio-environmental conditions to support overall wellbeing and psychosocial adaptation (Miller et al., 2021; Miller & Rasmussen, 2024). There is a growing evidence base on the effectiveness and acceptability of multilevel mental health and psychosocial (MHPSS) interventions (Goodkind et al., 2021; Moyano et al., 2024). Although conflict-related traumatic events present a significant risk to mental health, our result on the positive association between conflict-related traumatic experiences and social functioning highlight the potential usefulness of strategies fostering constructive meaning-making process following trauma (Pop et al., 2025) to support social functioning. Further, given the distinct nature of social functioning as a construct from mental health, promoting and protecting social functioning might require interventions with explicit social components such as strengthening social support and enhancing social capital. However, the majority of evidence-based MHPSS interventions primarily target intrapersonal processes, with minimal or no explicit focus on the social dimensions of wellbeing (Barbui et al., 2020; Schäfer et al., 2023). Moreover, even among those that include social components, limited number of studies evaluating their effectiveness typically rely on measures of psychological distress (Haroz et al., 2020; Ubels et al., 2022). Thus, there remains a critical gap in our knowledge about the effectiveness of existing MHPSS interventions for social functioning. Building on the growing evidence of the importance of social functioning among forcibly displaced communities (Kurt et al., 2025; McGrath et al., 2025), our scholarly observations from working with refugee communities and humanitarian workers across several low-resource settings, and the findings from this study, we call for a deliberate effort to understand and support social functioning as a central component of MHPSS programming, as it was originally envisioned.

In summary, this study extends the ecological model of distress to social functioning among refugees and asylum-seekers. The findings lend credence to social functioning as a distinct construct with the differential pathways from conflict-related traumatic experiences and post-displacement stressors. Recognizing social functioning as an integral component of the ecological model provides a strong conceptual foundation for advancing research and practice in MHPSS programming for refugees and asylum-seekers.

## Data availability

Data is not publicly available due to its sensitive nature. Anonymized data may be shared upon reasonable request to the corresponding author.

## Acknowledgments

We would like to thank each participant who entrusted us with their experiences. We acknowledge the contributions of our study partners, HOST International, SUAKA, and Universitas Gadjah Mada. Special thanks to Shraddha Kashyap, Mitra Khakbaz, Diah Tricesaria, Zico Pestalozzi, Yunizar Adiputera, and Shaila Tieken for their invaluable contributions to the study.

